# How should hospitals manage the backlog of patients awaiting surgery following the COVID-19 pandemic? A demand modelling simulation case study for carotid endarterectomy

**DOI:** 10.1101/2020.04.29.20085183

**Authors:** Guy Martin, Jonathan Clarke, Sheraz Markar, Alexander W. Carter, Sam Mason, Sanjay Purkayastha, Ara Darzi, James Kinross, on behalf of the PanSurg Collaborative

## Abstract

**Background:** The COVID-19 pandemic presents unparalleled challenges for the delivery of safe and effective care. In response, many health systems have chosen to restrict access to surgery and reallocate resources; the impact on the provision of surgical services has been profound, with huge numbers of patient now awaiting surgery at the risk of avoidable harm. The challenge now is how do hospitals transition from the current pandemic mode of operation back to business as usual, and ensure that all patients receive equitable, timely and high-quality surgical care during all phases of the public health crisis.

**Aims and Methods:** This case study takes carotid endarterectomy as a time-sensitive surgical procedure and simulates 400 compartmental demand modelling scenarios for managing surgical capacity in the UK for two years following the pandemic.

**Results:** A total of 7,69 patients will require carotid endarterectomy. In the worst-case scenario, if no additional capacity is provided on resumption of normal service, the waiting list may never be cleared, and no patient will receive surgery within the 2-week target; potentially leading to >1000 avoidable strokes. If surgical capacity is doubled after 1-month of resuming normal service, it will still take more than 6-months to clear the backlog, and 30.8% of patients will not undergo surgery within 2-weeks, with an average wait of 20.3 days for the proceeding 2 years.

**Conclusions:** This case study for carotid endarterectomy has shown that every healthcare system is going to have to make difficult decisions for balancing human and capital resources against the needs of patients. It has demonstrated that the timing and size of this effort will critically influence the ability of these systems to return to their baseline and continue to provide the highest quality care for all. The failure to sustainably increase surgical capacity early in the post-COVID-19 period will have significant long-term negative impacts on patients and is likely to result in avoidable harm.

## Introduction

The current COVID-19 pandemic is the biggest threat to health in living memory and presents unparalleled challenges for the delivery of safe and effective care at scale across all health systems, which have not been considered before. The pandemic has placed unprecedented pressures on acute healthcare resources and capacity; demand has overwhelmed even the most developed and well-resourced health systems[1], and difficult decisions regarding the rationing of care have now been made[2]. In response to the pandemic, many health systems have chosen to restrict access to surgical procedures and reallocate resources in order to create additional short-term hospital capacity[3–7]; the impact on the provision of surgical services has been profound with more than two million procedures cancelled to date in the UK at a cost of £3bn[8]. Patients will continue to develop a full range of pathologies as the pandemic progresses, continually contributing to the demand for post-pandemic surgical services. Ultimately, this will further exacerbate waiting times, cause preventable morbidity and mortality from surgical disease and adversely affect quality of life. As healthcare systems attempt to regain the initiative, policy makers and administrators must now determine how these systems adapt to meet their elective responsibilities and transition from the current crisis mode of operation back to ‘business as usual’. The challenge is to manage patient demand for surgery during all phases of the pandemic whilst ensuring equitable, timely and high-quality care for all. However, no models currently exist that allow policy makers to develop strategies for achieving this goal.

Stroke is one of the leading causes of death and long-term disability[9], and places a large burden on health systems, patients and their relatives. The principal cause of stroke is carotid territory ischaemia, with up to 15% of all cases attributable to significant carotid artery stenoses[10]. Carotid endarterectomy (CEA) is one of the most studied surgical procedures, and there is little controversy regarding its benefit in those with moderate to high-grade stenosis, with up to a 53% relative reduction in stroke risk compared to best medical therapy alone, and with optimal outcomes seen when surgery is performed within two weeks of symptoms[11,12]. Whilst there is some evidence for a decline in the number of procedures performed annually across the world[13,14], carotid endarterectomy remains a relatively common procedure with approximately 4,000 cases performed annually in the United Kingdom[15]. National bodies such as The Vascular Society for Great Britain and Ireland have issued guidance suggesting that surgeons should still try to operate on high risk patients during the pandemic, but that aggressive best medical therapy may be more appropriate[16]; the impact of COVID-19 on carotid surgery has been hugely significant. This study therefore takes CEA as a case study for an evidence-based time-sensitive surgical procedure, and then seeks to simulate a range of demand modelling scenarios for managing surgical capacity after the pandemic has concluded in order to return to business as usual within the United Kingdom’s National Health Service (NHS). These models provide valuable insights that are applicable to all surgical procedures and health systems.

## Methods

### Compartmental Demand Model

A compartmental model was constructed to represent a range of scenarios for increasing surgical capacity for CEA following total cessation of surgical services in response to the COVID-19 pandemic. There is no monthly variation in stroke rates[17], and so for the purposes of the model a constant weekly rate of demand for CEA was assumed based on the number of procedures performed for symptomatic carotid disease in the United Kingdom in 2018; equating to 74 cases per week[15]. In addition, it was assumed that at baseline demand matches system capacity.

*Figure 1* shows the components of the model used. Each week, a finite number of patients from an infinite pool of potential patients are assumed to require carotid endarterectomy. These patients are added to the list of patients awaiting surgery. Each week, a finite number of patients are taken from this list and undergo surgery. Patients are selected on a ‘first come, first served’ basis, and so the patients operated on each week are those that have waited longest for surgery. In each model it is assumed that no carotid endarterectomies are performed for three months (13 weeks) due to COVID-19, after which surgery resumes. Throughout this period, new patients continue to require carotid endarterectomies at a constant rate of 74 cases per week. In the model it was assumed that once deemed to require surgery, all patients remained eligible and survived to undergo their procedure irrespective of delay.

**Figure 1.**
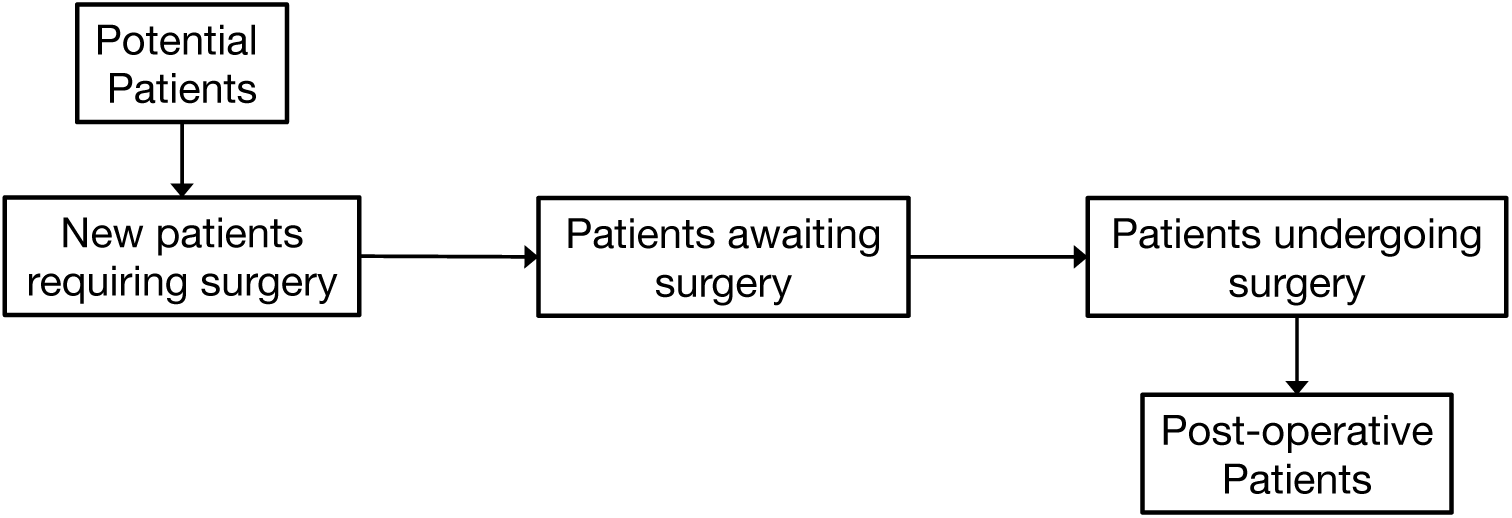
schematic of demand model components.

Different scenarios for the capacity to perform carotid endarterectomies were simulated subject to two distinct parameters; the time taken to return to baseline capacity (TTB) and the eventual additional capacity reached above pre-COVID-19 capacity (EC). Capacity is assumed to increase at a constant rate until EC is reached. Models were simulated for the two years after the initial cessation of surgical services for twenty different TTB values, ranging from 0 to 19 weeks, and 20 different EC values, ranging from 0% to 100% of baseline capacity. *Figure 2* illustrates three of the 400 potential capacity scenarios examined. Model 1 indicates an immediate return to pre-COVID-19 baseline capacity of 74 cases per week (TTB = 0, EC = 100), which thereafter remains constant. Model 2 indicates a return to pre-COVID-19 baseline capacity at 4 weeks, after which capacity continues to increase at the same rate to reach 20% above baseline capacity (TTB = 4, EC = 20). Model 3 indicates a slower return to baseline capacity, taking 8 weeks, after which capacity continues to increase to 40% above baseline capacity (TTB = 8, EC = 40). In each case once eventual capacity is reached, the number of procedures performed remains constant at this rate.

**Figure 2.**
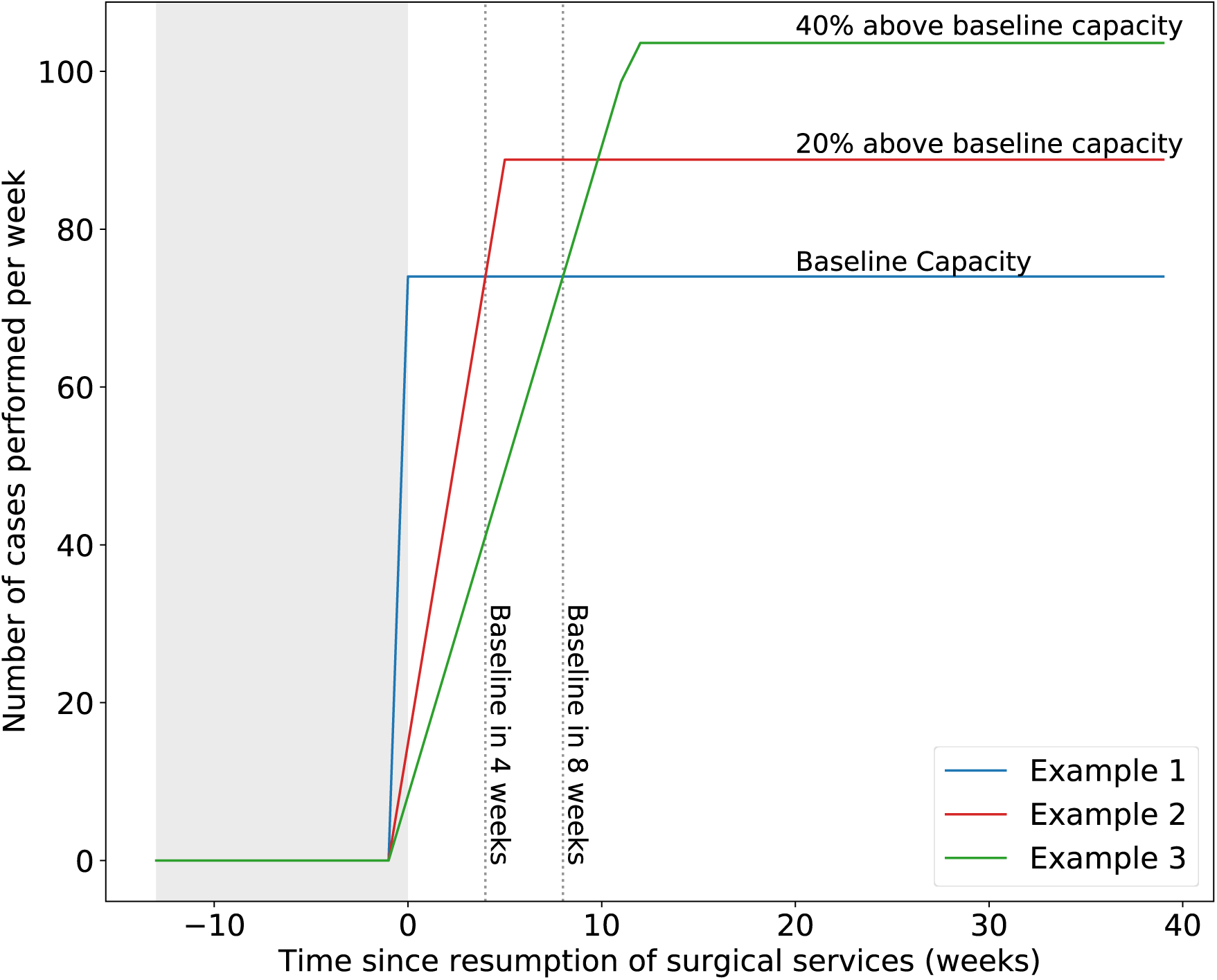
three illustrative examples of surgical capacity scenarios varying according to the time to return to pre-COVID-19 baseline capacity (TTB) and the eventual capacity reached (EC) following complete cessation of surgery for three months. Model 1) pre-COVID-19 capacity is restarted on day 1 and maintained. Model 2) pre-COVID-19 capacity is reached after 4 weeks and then increases to reach 20% above baseline. Model 3) pre-COVID-19 capacity is reached after 8 weeks and then increases to reach 40% above baseline.

The full mathematical model and code is available on request from the authors. All simulation, analysis and production of figures was conducted using Python V3.6.8 (Python Software Foundation, www.python.org) using the pandas, numpy, matplotlib and scikit.learn libraries.

### Outcome Measures

The primary outcome measure reported is the mean waiting time for patients newly requiring carotid endarterectomy in the two years from the start of the period of cessation of surgery. The time taken for the waiting list to be eliminated, and the proportion of patients waiting 2 weeks and 12 weeks for surgery are reported as secondary outcome measures. These were selected based on data derived from the most recent pooled analysis of 6,092 patients with 35,000 years of follow-up that shows maximal benefit from surgery when performed within 2 weeks of symptoms, and no benefit after a wait of more than 12 weeks[11]. Contour plots were produced using cubic interpolation of scatter points to represents scenarios of equal values with respect to the above outcome measures.

## Results

A total of 7,696 patients were assumed to require carotid endarterectomy in the two years following the start of cessation of surgical services. Where surgical capacity immediately returns to pre-COVID-19 capacity of 74 cases per week (Mode 1, *Figure 2*), surgical capacity equals surgical demand, and therefore waiting lists remain at a constant size of 962 cases after the resumption of surgery. The waiting list does not reduce, owing to new demand being equal to the number of cases performed each week.

*Figure 3* shows the performance of the 400 models simulated with respect to the average time waited for surgery, the time taken for the waiting list to be eliminated and the proportion of patients waiting more than 2 and 12 weeks for surgery is shown.

**Figure 3.**
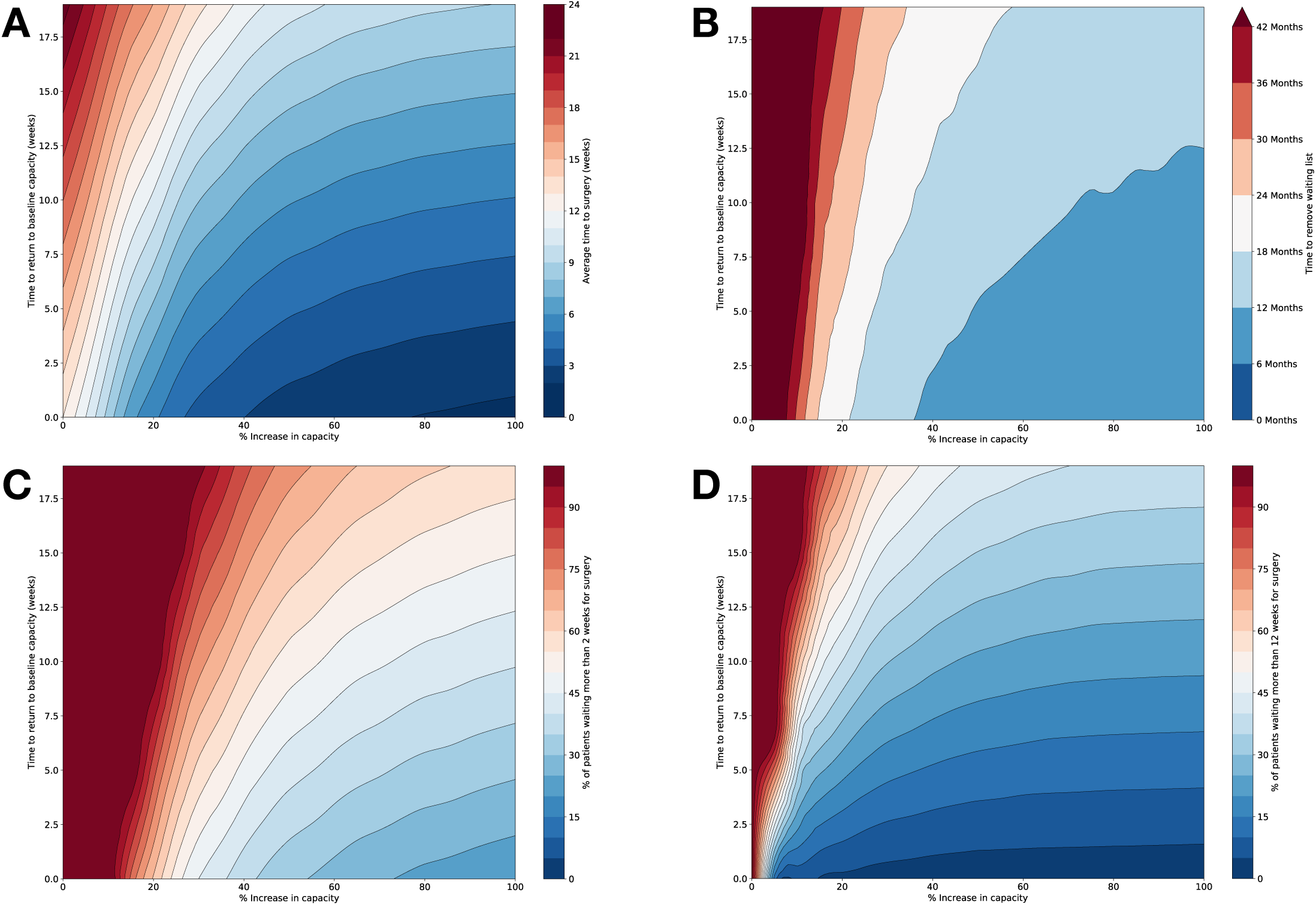
contour plots showing the performance of simulation models over varying times to return to baseline capacity (TTB) on the y-axis and the eventual additional capacity reached (EC) on the x-axis for patients requiring surgery within 2 years of the onset of the 3-month cessation of surgery. A) average wait time for patients to undergo surgery. B) time to clear waiting list and return to baseline. C) proportion of patients undergoing surgery within 2 weeks. D) proportion of patients undergoing surgery within 12 weeks.

*Figure 3A* shows that above an EC of 40%, the average time waited for surgery is primarily dictated by the TTB, while for EC of less than 20%, EC is more influential with respect to average waiting time than TTB. For a 20% increase in capacity above baseline, an average waiting time of 12 weeks can only be achieved if pre-COVID-19 baseline capacity is reached in fewer than 10 weeks. Where eventual additional capacity is double pre-COVID-19 baseline capacity, if TTB is 8 weeks, patients would on average wait 4 weeks for surgery.

*Figure 3B* shows the time taken for the additional waiting list to be eroded is more than two years where capacity is increased above pre-COVID-19 baseline capacity by less than 20%, unless capacity is returns to baseline in less than 3 weeks. For capacity increases of 10% or less, the additional waiting list is not eroded completely for at least three years. Even if capacity is doubled, unless baseline capacity is reached within three months, it would take at least a year to erode the additional waiting list as a result of cessation of surgical services. Under all scenarios, the additional waiting list persists for at least 6 months.

*Figure 3C* shows a 30% increase in capacity above the pre-COVID-19 baseline, coupled with a TTB of 0 weeks is required for more than 50% of patients to receive a carotid endarterectomy within 2 weeks in the two years following cessation of services. In scenarios where pre-COVID-19 baseline capacity is reached after two months following resumption of surgical services, capacity increases of 60% or more are required to ensure half of patients receive surgery in less than 2 weeks.

*Figure 3D* shows where capacity is increased to 20% or more above baseline capacity, 50% or more of patients receive carotid endarterectomies within 12 weeks. For capacity increases of less than 20%, the time taken to return to baseline capacity strongly influences the proportion of patients operated on within 12 weeks.

## Discussion

The cessation of the majority of surgery has had a profound impact on services and will undoubtedly lead to huge increases in patients awaiting intervention as the incidence of non-traumatic surgical pathology is unlikely to change as a result of the pandemic. Whilst the initial priority for policy makers and clinicians was rightly managing the acute phase of the pandemic, the next, and potentially far bigger challenge is determining how and when hospitals return to business as usual in what will be a post-pandemic new normal.

This demand modelling scenario has shown that following a 3-month cessation of carotid endarterectomy surgery, if no additional capacity is provided on resumption of normal service the waiting list will not be cleared for over 2 years, and the vast majority of patients would wait >12 weeks for surgery at which point it is of no benefit; exposing them to double the risk of stroke and resulting in the region of 350–1100 additional strokes[11]. Even if surgical capacity is doubled after 1-month of resuming normal service it will still take over 6-months to clear the waiting list backlog, 30.8% of patients will not undergo surgery within the 2-week target, and 11.5% will wait more than 12-weeks with an average wait to surgery of 20.3 days for the proceeding 2 years; resulting in the region of 100–350 additional strokes. It should also be noted that this modelling technique is not specific to CEA, with delays in surgery from a post-COVID-19 backlog in cases likely to have a direct impact on patients across a wide range of diagnoses. Given these sobering figures hospitals must now start to look forwards and plan for how they can and should manage capacity to ensure they provide equitable, timely and high-quality care for all in the post-COVID-19 recovery phase.

Increasing capacity to deal with excess demand is not straight forward, with multiple factors influencing the ability of hospitals to increase the number of surgical procedures performed and clear the post-COVID-19 backlog.

External factors may include unpredictable changes in the rate of new stroke diagnoses due to a reduction in the number of patients presenting with acute symptoms during the pandemic; changes which have already been reported across a range of diagnoses including in stroke[18,19]. This phenomenon will be of even greater importance in diseases with more complex referral and treatment patterns such as gastrointestinal cancer where the majority of patients move between primary care, screening services, diagnostics and hospitals prior to receiving surgery. Additionally, there are going to be inevitable further spikes in COVID-19 cases until either a vaccine is developed or herd immunity is achieved[20]. This will lead to further fluctuations in surgical capacity and successive shut downs over time; it is therefore it is important to meaningfully catch up when excess capacity is possible, and more elegantly decrease surgical capacity without a total shutdown during future spikes to ensure we are not once again starting from a standstill.

In addition to external factors, multiple internal considerations will also influence the ability to increase capacity and catch up with a surgical backlog as highlighted in current capacity planning models[21]. Surgical teams do not operate in isolation, and critical care capacity is vital to support a wide range of surgical procedures including CEA. The COVID-19 pandemic has led to a huge increase in demand for critical care capacity that will likely continue into the medium- and long-term[22,23], and even before the crisis critical care units typically had occupancy rates of around 80–85%[24,25] with significant variability in the availability of critical care capacity in relation to surgical activity[26]. Surgical teams can therefore not depend on predictable levels of critical care availability to support an increase in capacity and so will need to plan accordingly or modify peri-operative practice. It is not just critical care capacity that may limit the ability to rapidly expand surgical services. The NHS in England typically runs at general bed occupancy rates of 90-95%[27], and therefore its ability to rapidly expand surgical numbers is severely limited without seeking alternative infrastructure. Potential options could include the use of the private healthcare sector or transitioning temporary facilities such as the NHS Nightingale Hospitals that were built in response to the COVID-19 crisis to more permanent facilities geared towards reducing waiting lists by providing additional capacity. The most important resource for clearing surgical waiting lists is staff though. Burnout is an increasingly recognised phenomena across all healthcare workers, and especially in those delivering surgical services[28,29]. Compounding this will be the knock on effects of the huge increases in work intensity, the redeployment of staff across different services and the consequences of the sobering 20% COVID-19 infection rates in healthcare staff[30]. Even if physical capacity is found, healthcare providers must carefully look after, protect and support their staff to ensure that the post-COVID-19 response and return to business as usual is both successful and sustainable; healthcare workers are human, and have limits.

The model presented does have limitations. It assumes all patients remain eligible for intervention and survive to undergo their procedure irrespective of delay. Within the CEA cohort there is likely to be in the region of a 10–20% annual dropout rate due to death or further stroke that makes someone ineligible for surgery, with the greatest risk seen in the first three months[31,32]. This would act to reduce the number of patients requiring intervention, and thus the time to return to baseline, but the impact will be minimal; importantly, the model represents the worst-case scenario for which health systems should plan for, and act on. In addition, this is a generalised model based on national figures. As such, there is a requirement for individual organisations to take into account local factors such as the direct disruption caused by COVID-19, competing demands of surgical and supporting services, and wider infrastructure and staffing resource constraints when designing, modelling and implementing local recovery plans.

To further refine the models and identify contextual solutions, discrete event simulations could be employed to incorporate probabilistic payoffs on decisions around additional factors such as mortality rates, the social acceptability of delays and impact of local and national prioritisation and planning decisions. It is also important to assess the financial and human capital implications of seeking to increase surgical capacity. To address the avoidable burden of stroke, the health implications and costs of these policy decisions could be expressed as incremental gains or losses in quality adjusted life years (QALY). We recommend that future analyses incorporate these cost indicators, in addition to risk-based decision aids to help health systems allocate their finite resources to maximise patient benefit.

The cessation of the majority of global elective surgical services during the Covid-19 pandemic has had a profound impact on patients and healthcare providers. This case study for carotid endarterectomy has shown that every healthcare system is going to have to make difficult decisions when balancing human and capital resources against the needs of patients. It has also demonstrated that the timing and size of this effort will critically influence the ability of these systems to return to their baseline and continue to provide the highest quality care for all. The failure to sustainably increase surgical capacity early in the post-COVID-19 era will have significant long-term negative impacts on patients and is likely to result in avoidable harm.

## Data Availability

All original code and data is available from the corresponding author upon request

